# Digital adherence technologies to improve tuberculosis treatment outcomes: a cluster-randomised superiority trial

**DOI:** 10.1101/2023.01.25.23285001

**Authors:** Xiaoqiu Liu, Jennifer Thompson, Haiyan Dong, Sedona Sweeney, Xue Li, Yanli Yuan, Xiaomeng Wang, Wangrui He, Bruce Thomas, Caihong Xu, Dongmei Hu, Anna Vassall, Shitong Huan, Hui Zhang, Shiwen Jiang, Katherine Fielding, Yanlin Zhao

## Abstract

**Background:** Drug-sensitive tuberculosis treatment is for six months; adherence problems are common. Digital adherence technologies may improve outcomes.

**Methods:** In a cluster-randomised trial, 24 counties/districts in China were randomised (1:1) to two groups. Patients received: a medication monitor for daily drug-dosing reminders and health care worker monthly adherence monitoring with management of patients with poor adherence (intervention); or routine care (control; silent-mode monitor measured adherence). Adults with GeneXpert-positive drug-sensitive tuberculosis were enrolled and followed-up with sputum (solid culture) at 12 and 18 months. The objective was to assess whether digital adherence technologies combined with health care worker support for patients struggling with adherence improves treatment outcomes and reduces recurrence. The primary composite unfavourable outcome was death/lost-to-follow-up/failure on treatment or recurrence by 18 months from treatment start. Secondary outcomes included adherence. 12 clusters/group (125 patients/cluster), unfavourable outcome of 18% in control, coefficient of variation 0.3, gave 85% power for a 40% reduction in outcome. Analysis accounted for study design with multiple imputation for the primary outcome. Only the independent endpoints review committee who assessed endpoint data for some participants were masked to study group. The trial was registered at Current Controlled Trials (ISRCTN35812455).

**Findings:** From Jan2017-Mar2019, 3075 patients were enrolled and 2686 (87%) contributed to the primary outcome. Post-randomisation two intervention clusters were merged. Overall 71% were male, median age 44 years. Of 433 unfavourable outcomes, 289(67%) were treatment lost-to-follow-up, 42(10%) recurrence. The intervention had no impact on unfavourable outcome (adjusted risk ratio 1.01, 95% confidence interval 0.73-1.4) and other treatment outcomes. Treatment non-adherence was reduced by 60-65%.

**Interpretation:** Our medication monitor intervention did reduce non-adherence but had no impact on the unfavourable outcome which included lost-to-follow-up and recurrence. There was a failure to change management following identification of non-adherence at monthly reviews. Recurrence was rare and measurement may have been limited due to programmatic conditions and using solid culture.

**Funding:** Bill & Melinda Gates Foundation

**Research in context:** *Evidence before this study:* Prior to the start of the study we searched Medline and Embase (December 2015) using search terms (digital pill box* OR smart pill box* OR SMS OR text messag*) AND TB or tuberculosis. We found one systematic review assessing the effect of mobile phone text messaging on treatment adherence used as a proxy for treatment outcomes and development of drug resistance. Four studies (three observational and one randomised trial) were included, meta-analysis was not conducted, and authors concluded mixed findings for the effectiveness of text messaging to promote adherence. Our previous study in China, published in 2015, reported improved adherence to TB treatment with text messaging and/or smart pill box reminders. The study was not powered for treatment outcomes. Since then two studies have reported improved TB outcomes. A study conducted in Kenya assessed weekly motivational messages, daily text reminders, a USSD platform for patients to confirm daily adherence followed by SMS and calls from the research team for patients who had not confirmed adherence and clinic notification of patients with no confirmation for more than 2 days. The intervention reduced unsuccessful outcome by 68%, entirely through reducing loss to follow-up. The second study was a stepped-wedge trial from Uganda assessing a text messaging based intervention, where patients received daily text dosing reminders and were asked to confirm a dose taken using a toll-free number. Adherence data were reviewed at clinics visits every two weeks or monthly resulted in differentiated management. The authors showed improved successful treatment outcomes, though among a per-protocol population (97% and 52% of the populations in the control and intervention phases) who enrolled onto the intervention within the first two months of treatment. A recent systematic review in 2022 reported variable effects of digital adherence technologies on treatment outcomes.

*Added value of this study:* This is the first trial to report the impact of a digital adherence technology intervention (smart pill box reminder, monthly review of adherence data and differentiated care for those where lack of pill box opening, as a proxy for adherence, was a problem) on a composite unfavourable endpoint of poor treatment outcome or subsequent retreatment including culture-confirmed recurrence, among drug-sensitive patients. The study found that monthly review of adherence data was not adequate to influence poor treatment outcomes, in particular losses to follow-up, or recurrence. There was a failure to change management following identification of non-adherence at the monthly reviews. We did demonstrate, however, a reduction in non-adherence in the intervention versus standard of care, similar to our previous study, indicating improved quality of treatment with the smart pill box intervention.

*Implications of all the available evidence:* Currently there is no strong evidence that digital adherence technology interventions improve health outcomes, including treatment recurrence. More frequent review of adherence data with a streamlined approach for identifying patients with adherence issues and escalating supportive management of these patients, may be key for improving outcomes.

## Introduction

An estimated 10 million people fell ill with tuberculosis in 2019(1). Recent declines in tuberculosis incidence and deaths have been observed, however these are unlikely to be fast enough to reach reduction milestones for 2030. China is among eight countries contributing two-thirds of the global tuberculosis total, though 2015-2019 has seen a reduction in tuberculosis incidence and total number of deaths of 10% and 22%, respectively(1).

National guidelines recommend daily fixed-dose combination to treat drug-sensitive tuberculosis of two months of isoniazid, rifampin, pyrazinamide and ethambutol, followed by four months of isoniazid, rifampin (2HRZE/4HR). High levels of treatment adherence are considered important for cure and reducing recurrence(2, 3).

A major component of the Directly Observed Treatment Short-course strategy, introduced in China in 1992 and covering the whole country by 2005, is Directly Observed Treatment (DOT) to help improve medication adherence. A systematic review of treatment support using studies from China showed, however, only 20% of patients had DOT by a health professional and over half were self-administering treatment(4). Despite this, China’s treatment success for new and relapse drug-sensitive tuberculosis was reported to be 94%(1).

Digital adherence technologies including short message service (SMS) and electronic pill boxes, which support patients in their adherence, have the potential of enhancing patient care through improving interactions between patients and health care providers, and increasing treatment adherence and successful treatment outcomes(5, 6). WHO’s drug-sensitive tuberculosis updated treatment guidelines, made a conditional recommendation with very low certainty of evidence for tracers (such as mobile phone SMS) and/or digital medication monitors may being offered to tuberculosis patients(7).

Two studies have demonstrated improved treatment outcomes: through reducing loss-to-follow-up using SMS and an Unstructured Supplementary Service Data (USSD)-based intervention in Kenya and improving treatment success in a stepped wedge trial in Uganda, among a subset of patients exposed to the SMS-style intervention(8, 9).

We report on a cluster-randomised trial to evaluate the impact of a daily reminder medication monitor, monthly review of adherence data by health care provider with patient, and differentiated care for those with adherence issues, on unfavourable treatment and adherence outcomes.

## Methods

### Study design

In this pragmatic, cluster-randomized trial in four prefectures of China, geographical areas (clusters) served by a TB dispensary or designated hospital were the unit of randomisation(10). Patients were assigned to the cluster according to the TB dispensary or designated hospital where they received their tuberculosis treatment.

The trial was approved by Institutional Review Board of the Chinese Center for Disease Control and Prevention and the London School of Hygiene & Tropical Medicine Ethics Committee.

### Cluster and participant inclusion

Clusters had >300 pulmonary tuberculosis patients in 2014, access to GeneXpert and culture diagnosis, TB services supplied by a TB designated hospital or dispensary, and implementing a daily chemotherapy scheme. Consecutive patients were enrolled if they had pulmonary, GeneXpert-positive and rifampicin-sensitive tuberculosis, on daily fixed-dose combination treatment, able to attend follow-up visits 12 and 18 months after treatment start. Participants provided written informed consent to join the trial.

### Randomisation and interventions

Constrained randomisation was used to allocate 24 clusters (ratio 1:1) to intervention or control (routine care) group balanced for prefecture (by prefecture, difference by group was at most one), health setting type (hospital/dispensary; seven hospital and five dispensary in each group), area (urban/rural; by area, difference by group was at most one) and sputum smear-positive tuberculosis notifications in 2015 (difference in average notifications by group was at most 10 cases). Randomisation was conducted by the trial statistician using Stata (version 14). Post-randomisation it was identified that two intervention clusters used the same dispensary, so were combined into one cluster.

Cluster-randomisation was justified to reduce contamination between groups and for logistical convenience: the intervention required changes to the delivery of care raising concerns that individual randomisation would lead staff to change their behaviour towards control patients and patients to discuss care with one another.

### Procedures

In both groups, patients were given a daily tuberculosis treatment regimen of 2HRZE/4HR and a medication event reminder monitor (MERM) to store medication. The MERM recorded the dates and times that it was opened for more than 2 seconds for patients to take their medication, and recorded a “heartbeat” daily to indicate it was working.

In intervention clusters the MERM gave an audio and visual reminder to take medication (daily, three times within five minutes) and attend monthly clinic visits. At clinic visits, the doctor could display monitor openings in the last month on their computer in order to discuss adherence with the patient. Intensive management was initiated the first time non-adherence was between 20%-50% in the previous month; township doctors would be asked to visit patients every two weeks and village doctors every week. The second month non-adherence reached this level or the first month non-adherence was >50%, patients were switched to DOT with medical staff observing therapy administration.

Patients in control clusters had the reminder functions on the MERM disabled. Managing doctors could not review monitor openings at clinic visits, but data were collected for the trial. In consultation with the doctor, patients chose whether to take medication under direct observation by a healthcare worker, family member, or through self-administration.

Patients attended monthly routine clinic visits for the six months of treatment. Routine sputum collection for smear microscopy was conducted at months 2, 5 and end of treatment. For the latter an additional sputum was collected for solid culture (using routine laboratories) and a chest radiograph conducted. Patients were telephoned 9 and 15 months after the start of treatment and self-reported retreatment for tuberculosis. At 12 and 18 months after the start of treatment, patients attended the clinic to give sputum for culture, have a chest radiograph, and self-report tuberculosis retreatment.

Missed doses were measured by days no opening was recorded on the MERM restricted to days patients reported using the monitor and it had a heartbeat.

### Outcomes

The primary outcome was a composite unfavourable outcome defined as poor treatment outcome (treatment failure, died, developed multidrug resistant tuberculosis, lost-to-follow-up, or stopping treatment due to an adverse reaction or refusal of treatment) or tuberculosis recurrence within 18 months of starting treatment. Recurrence was defined as a single positive culture, chest radiograph satisfying the case definition for new active tuberculosis, or self-report of retreatment. Recurrences identified by chest radiograph or self-report were reviewed by an independent endpoint review committee masked to group.

Secondary outcomes were components of the primary composite outcome (poor treatment outcome; lost-to-follow-up during treatment; and poor outcome or recurrence in 12 months), time to recurrence in those who had cured or completed treatment, and two-month smear conversion among those smear-positive at the start of treatment. Secondary outcomes for adherence were percentage of months in which patients missed at least 20% of doses, percentage of overall doses missed, and visits attended on schedule. Process measures included inability to use the fixed dose combination treatment, number of visits by township and village doctor, MERM malfunctions and withdrawals from using the MERM. In the intervention group we also summarised the number of times the alarm sounded each day, and change of management due to non-adherence. Visits were defined as being attended on schedule if the number of days between visits was the same or less than the number of days of medication given to patients at their previous visit.

### Statistical analysis

In original sample size calculations, 12 clusters per group and a harmonic mean 125 patients per cluster gave 85% power to detect a 40% reduction in unfavourable outcome at the 5% level assuming 18% risk of unfavourable outcome in the control group, 5% lost-to-follow-up, and a coefficient of variation of outcome of 0.3. Recalculation after two clusters were combined, a harmonic mean 108 patients per cluster due to slower than expected recruitment, and allowing 10% lost-to-follow up gave 83% power.

The intention-to-treat population was defined as participants enrolled into the trial, excluding those satisfying a limited number of post-enrolment exclusions (participants who stopped taking the fixed-dose combination treatment within the first 1 month due to an adverse reaction; permanently stopped their treatment management model within the first 1 month due to travel or hospitalisation; treatment extended due to updated diagnosis of TB pleurisy, or trachea and/or bronchus TB; diagnosis of drug resistance due to non-rifampin drug-resistance), and participants with a change of diagnosis confirmed by the endpoint review committee. The per-protocol population further excluded participants who withdrew early from use of the MERM regardless of the reason given.

All analyses were conducted in Stata (version 15) using the clan command. We use a cluster-level analysis. Our primary estimand was a risk ratio, calculated using the logarithm of the cluster proportions: this estimates the ratio of geometric means of the cluster-level risks in each group, so all percentages reported are geometric means. Our primary analysis was adjusted for age, sex, occupation, migrant status, distance to clinic, education level, household expenditure, and smear result at treatment initiation using the two stage approach(11).

For the primary outcome, the primary analysis was based on multiple imputation using the intention-to-treat population. Multiple imputation due to missing composite outcome was applied with 25 imputations. See the supplement for further statistical methods. Complete case analyses for the intention-to-treat and per-protocol populations were also conducted. All secondary outcomes were analysed using complete cases only. For the primary outcome using the intention-to-treat population, pre-specified subgroup complete case analyses were conducted for urban/rural, clinic type, age, literacy level, sex, and household expenditure, and post-hoc subgroup analyses were conducted for smear status and GeneXpert cycle threshold.

The trial is registered with Current Controlled Trials (identifier ISRCTN35812455).

## Results

Of 23 clusters enrolled (seven Ganzhou, six Hangzhou, three Jilin, seven in Wenzhou), 14 treated patients in TB hospitals and 17 were in urban areas.

Patients were enrolled between 26 January 2017 and 3 April 2019. In the control group, 8179 patients were screened and 6743 excluded due to ineligibility, largely due to patients requiring treatment for >6 months (3249) or negative or rifampicin-resistant GeneXpert result (1993), and 48 did not provide consent. This was similar for the intervention group: 7078 patients were screened with 5752 excluded due to ineligibility (2663 required treatment >6 months and 1786 has a negative GeneXpert or rifampicin resistance), and 28 did not provide consent. This resulted in 1388 and 1298 patients in the control and intervention groups, respectively (figure 1).

**Figure 1:**
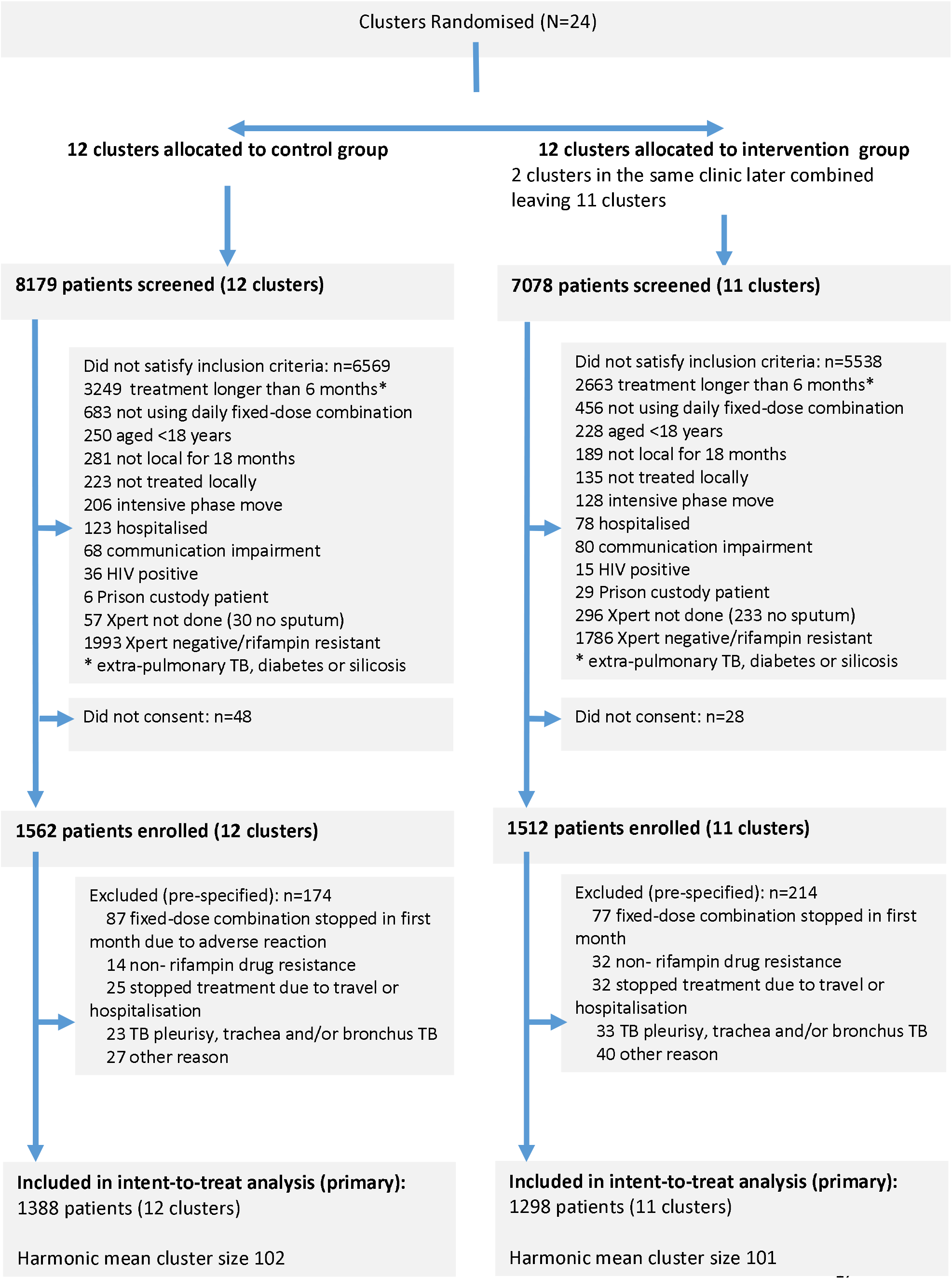
CONSORT diagram

Overall, 1909 (71%) were male, median age was 44 years (interquartile range [IQR] 29-58), and 1675 (62%) were smear-positive (Tables 1 and S1). Employment as a farmer was more common in the control (798/1387 [58%]) than the intervention group (585/1282 [45%]), and patients in the control group were more likely to be local residents (1088 [78%] versus 883 [68%] in the intervention group).

**Table 1:** Baseline characteristics of trial clusters and participants

| Characteristic |  | Control | Intervention |
| --- | --- | --- | --- |
| Cluster level covariates |  | N=12 | N=11 |
| Prefectures | Ganzhou | 4 (33%) | 3 (27%) |
|  | Hangzhou | 3 (25%) | 3 (27%) |
|  | Jilin | 2 (17%) | 1 (9%) |
|  | Wenzhou | 3 (25%) | 4 (36%) |
| Type n(%) | Hospital | 7 (58%) | 7 (64%) |
|  | TB dispensary | 5 (42%) | 4 (36%) |
| Urban n(%) |  | 9 (75%) | 8 (73%) |
| Number of sputum smear positive TB cases notified in 2015 median(IQR) |  | 148 (93.5-235) | 145 (113-182) |
| Individual level covariates |  | N = 1388 | N=1298 |
| Male n(%) |  | 989 (71%) | 920 (71%) |
| Age median (IQR), years |  | 45 (29 - 59) | 42 (29 - 57) |
| Marital status n(%) | Single | 299 (22%) | 272 (21%) |
|  | First marriage | 990 (71%) | 965 (74%) |
|  | Other | 99 (7%) | 61 (5%) |
| Employment n/N(%) | Unemployed (including students and retired) | 261/1387 (19%) | 194/1282 (15%) |
|  | Farmer | 798/1387 (58%) | 585/1282 (45%) |
|  | Other | 328 /1387 (24%) | 503/1282 (39%) |
| Education level n(%) | Illiterate | 126 (9%) | 99 (8%) |
|  | Primary | 404 (29%) | 359 (28%) |
|  | Junior middle | 494 (36%) | 500 (39 %) |
|  | High | 205 (15%) | 218 (17%) |
|  | University | 159 (11%) | 122 (9%) |
| Local resident n(%) |  | 1088 (78%) | 883 (68%) |
| Household expenditure, CNY, n(%) | < 1000 | 182 (13%) | 77 (6%) |
|  | 1001 - 3000 | 644 (46%) | 741 (57%) |
|  | ≥3001 | 562 (40%) | 480 (37%) |
| Smear-positive n(%) |  | 858 (62%) | 817 (63%) |

IQR: Interquartile range; CNY Chinese yuan renminbi
Percentages are overall, ignoring clustering

components of the unfavourable outcome are summarised in table S2. Overall 6·3% (88/1388) and 4·6% (60/1298) had missing unfavourable outcome at 18 months in the control and intervention groups, respectively. Using multiple imputation for missing outcomes, unfavourable outcomes occurred in 16% of patients in the control group (geometric mean of cluster-level percent [GM] based on mean of 25 multiple imputations; 239/1388 patients) and 16% of patients in the intervention group (224/1298 patients). There was no evidence of a difference in the risk of unfavourable outcomes between groups (p=0·95, adjusted risk ratio[aRR] 1·01, 95% confidence interval [CI] 0·73,1·40, adjusted risk difference [RD] 0·7% 95% CI -4·5%, 5·9%). Results were similar in unadjusted, complete case, and per-protocol analysis (Tables 2, S3 and S4).

**Table 2:** Primary outcome and secondary outcomes, excluding adherence

| Outcome | Control<sup>a</sup><br>N=1388 | Intervention<sup>a</sup><br>N=1298 | Unadjusted Risk<br>Ratio (95% CI) | Adjusted Risk<br>Ratio (95% CI) |
| --- | --- | --- | --- | --- |
| **Primary:** |  |  |  |  |
| unfavourable<br>outcome |  |  |  |  |
| Poor treatment<br>outcome or recurrence<br>in 18 months n(%)<sup>b</sup> | 239/1388 (16%) | 224/1298 (16%) | 0.99 (0.66, 1.48)<br>P=0.96 | 1.01 (0.73, 1.40)<br>P=0.95 |
| **Secondary:** |  |  |  |  |
| Poor treatment<br>outcome n/total n(%) | 203/1350 (14%) | 188/1283 (13%) | 0.94 (0.61, 1.45) | 0.97 (0.69, 1.35) |
| Lost-to-follow-up<br>during treatment<br>n/total n(%) | 156/1350 (10%) | 133/1283 (9%) | 0.90 (0.54, 1.50) | 0.92 (0.59, 1.44) |
| Poor outcome or<br>recurrence in 12<br>months n/total n(%) | 215/1322 (15%) | 204/1240 (15%) | 1.00 (0.68, 1.49) | 1.03 (0.75, 1.41) |
| Time to recurrence<br>n/total n (rate per 100<br>person-years)<sup>c</sup> | 14/1147 (1.0) | 28/1095 (1.6) | 1.60 (0.75, 3.43) | - |
| Two month smear<br>conversion to negative<br>n/total n(%)<sup>d</sup> | 689/759 (89%) | 639/729 (89%) | 1.00 (0.91, 1.08) | 0.99 (0.92, 1.06) |
| All using intention to treat population. All comparisons are intervention versus control |  |  |  |  |
| <sup>a</sup> n/N ignoring cluster. Percentages are the geometric mean of the cluster-level risk or rate of an event |  |  |  |  |
| <sup>b</sup> Calculated using multiple imputation. Values shown by group are the imputation-mean of total number of events and geometric mean of cluster-level proportion of events. All other outcomes used complete cases |  |  |  |  |
| <sup>c</sup> Rate difference reported between the groups, in those with a good treatment outcome. Adjusted analysis not completed due to small number of events |  |  |  |  |
| <sup>d</sup> In those with a positive smear at the start of treatment |  |  |  |  |

Prespecified subgroup analysis did not reveal substantial differences in the intervention effect by clinic type, patient age, literacy, gender, or household expenditure (Figure 2). There was a suggestion that the intervention was harmful in urban clusters (unfavourable outcomes in 47/326[GM 11%] SoC versus 62/330[GM 21%] intervention; aRR 1·74 [1·02,2·98]). Unfavourable outcomes had a coefficient of variation of 0·3 and 0·4 in the control and intervention groups, respectively.

**Figure 2:**
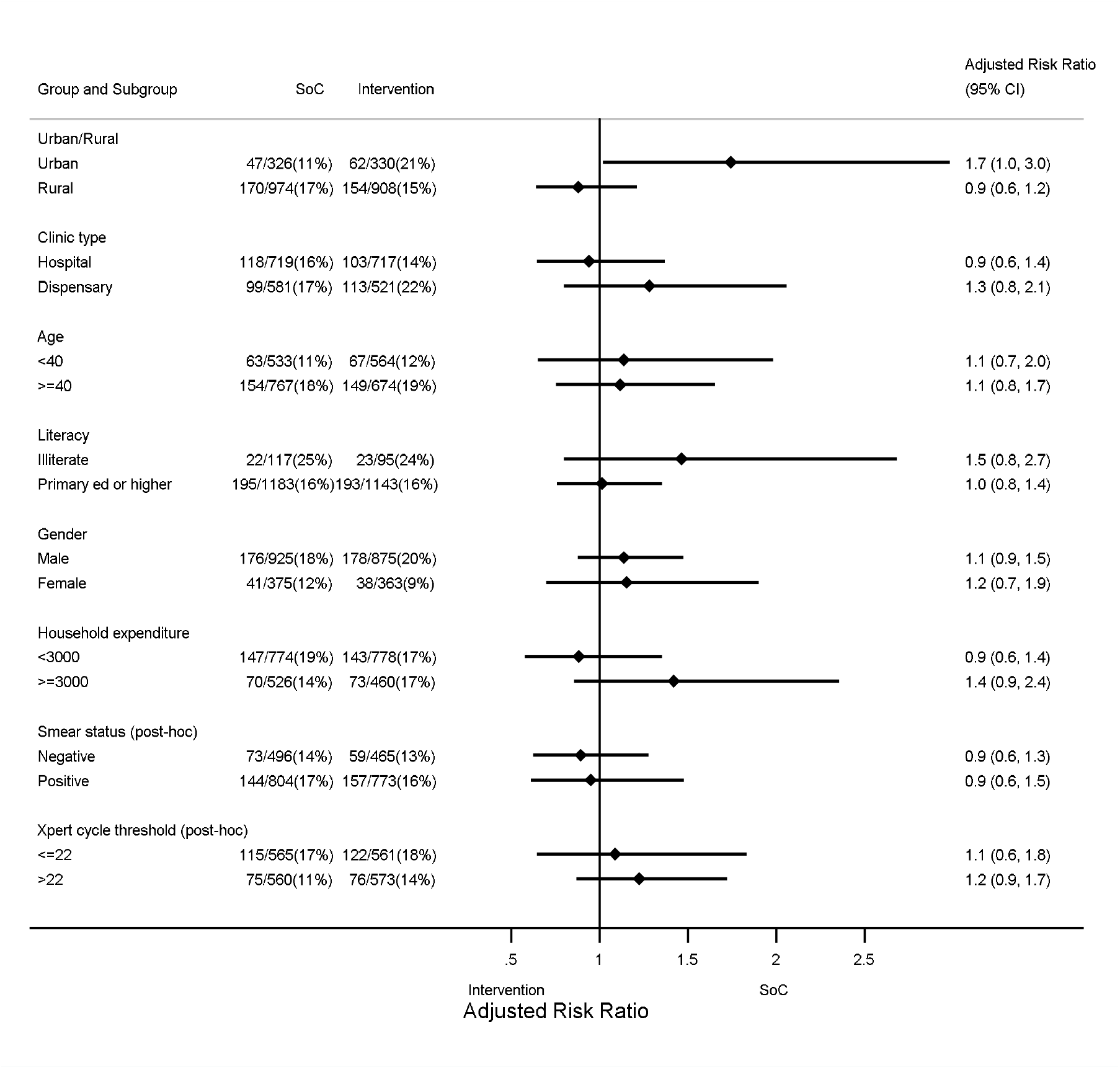
Subgroup analyses of composite outcome of poor treatment outcome and recurrence (primary outcome) using intention-to-treat population

Most unfavourable outcomes were due to poor treatment outcomes, which were similar between the groups (203/1350 [GM 14%] control versus 188/1283 [GM 13%] intervention; aRR=0·97 [0·69,1·35]), and most of these were due to patients becoming lost-to-follow-up on treatment (156/1350 [GM 10%] control versus 133/1283 [GM 9%] intervention; aRR=0·92[0·59,1·44]). In those with a successful treatment outcome, recurrence rates were similar by group (1·0 per 100 person-years control versus 1·6 per 100 person-years intervention; unadjusted rate ratio=1·60[0·75,3·43]). See tables 2 and S2.

Patients in the intervention group were 65% less likely to miss more than 20% of doses in a month (GM 2·7/6 months per person[46%] control versus 0·9/6[16%]; aRR=0·36[0·27,0·50]; table 3), and missed 57% fewer doses (GM 42/160 per person[28%] control versus 16/160[11%] aRR=0·43[0·34,0·53]).

**Table 3:** Secondary outcomes of medication adherence and process measures

| Outcome | Control | Intervention | Unadjusted group comparison (95% CI) | Adjusted group comparison (95% CI) |
| --- | --- | --- | --- | --- |
| **Adherence<sup>a</sup>** |  |  |  |  |
| Months in which patients missed >20% of doses / months of treatment per person (%) | 2.7/6.0 (46%) | 0.9/6.0 (16%) | 0.34 (0.24, 0.49) | 0.36 (0.27, 0.50) |
| Doses missed / doses expected per person (%) | 42/160 (27%) | 16/160 (11%) | 0.40 (0.31, 0.53) | 0.43 (0.34, 0.53) |
| Late or missed clinic visits / visits per person (%) | 2.6/5.0 (51%) | 2.5/5.0 (49%) | 0.96 (0.85, 1.08) | 0.97 (0.87, 1.08) |
| **Process measures** |  |  |  |  |
| Medication monitor errors days/treatment months (rate per treatment-month) | 2429/7097 (0.2) | 2812/6900 (0.4) | 1.77 (0.88, 3.56) | 1.75 (0.91, 3.36) |
| Quick openings n/treatment months (rate per treatment-month) | 48289/6987 (6.7) | 62309/6771 (9.0) | 1.33 (1.07, 1.66) | 1.30 (1.06, 1.60) |
| Withdrawals from use of MERM n/total n (%) | 105/ 1388 (6%) | 106/ 1298 (8%) | 1.27 (0.70, 2.33) | 1.26 (0.72, 2.23) |
| Unable to use FDC | 79/1388 (5%) | 46/1298 (4%) | 0.72 (0.33, 1.55) | 0.75 (0.36, 1.58) |
| Visits from township/village doctor per month |  |  |  |  |
| N (included in analysis) | 1247 | 1132 |  |  |
| Mean (sd)<sup>b</sup> | 2.5 (0.8) | 2.2 (0.8) | -0.2 (-1.0, +0.5) | -0.3 (-0.9, +0.4) |

<sup>a</sup> Adherence outcomes summarised by patient, taking the arithmetic mean within cluster, then the geometric mean between clusters.; <sup>b</sup> Before any change in management in the intervention arm. Group comparison is difference in means. All comparisons are intervention versus control
CI confidence interval; MERM medication event reminder monitor; FDC fixed dose combination; sd standard deviation

Post-hoc sensitivity analyses for the primary outcome and secondary adherence outcomes were consistent with the primary analyses (Tables S4, S5 and S6)

Most patients in the control group reported self-administering treatment (774/1156), and only 114/1156 were supervised by a health care worker (Figure S2).

Before any change in patient management in the intervention group, patients in both groups had similar contact with the township and village doctor between clinic visits (mean 2·5 visits control versus 2·2 visits intervention, adjusted mean difference=-0·3[-0·9,+0·4]). In the intervention group, intensive management was required by 196/1261 patients and was reported as received by 156/190. However, after patients’ switched to intensive management, there was no reported increase in township or village doctor contact (mean 1·8 visits in the preceding month in the intervention group Switching to DOT was required by 100/1261 patients and was reported as received by 53/99 (Table S7).

## Discussion

In this large cluster-randomised trial of 2686 drug-sensitive tuberculosis patients, the digital adherence technology intervention had no impact on reducing the risk of unfavourable outcome, poor end of treatment outcome or lost-to-follow-up during treatment. Recurrence was rare with a 12-month risk of 1·9% following the end of treatment among those who had not met poor end of treatment outcome. Non-adherence was reduced in the intervention versus control group by 60-65%, depending on the metric used, and which was a greater reduction compared with our previous study(6). A recent systematic review of digital adherence technologies to improve tuberculosis treatment outcomes reported intervention effects in different directions(12).

The intervention may have had no impact on lost to follow-up, the main component of the unfavourable outcome (67%; 289/433), for two reasons: lack of timely adherence data and failure to change management following identification of non-adherence at monthly reviews. Rather than monthly adherence assessment, a more frequent review of adherence data by health care workers and initiation of intensive management to assist patients who are struggling with adherence are likely needed to reduce loss-to-follow-up. The “Keheala” intervention assessed weekly motivational messages, daily SMS-reminders, a USSD platform for patients to confirm daily adherence with follow-up by the research team for patients who had not confirmed adherence and clinic notification of patients with no confirmation for >2 days. The intervention reduced unsuccessful outcome by 68%, entirely through reducing loss-to-follow-up(8). A stepped-wedge trial from Uganda demonstrated improved successful outcomes, including not being lost-to-follow-up, only among a per-protocol population who enrolled onto the intervention within the first two months of treatment(9). The study assessed a SMS-based intervention (99DOTS), whereby patients received daily SMS dosing reminders and were asked to confirm a dose taken using a toll-free number, as well as a weekly automated interactive voice response check-in. Review of adherence data at visits every two weeks or monthly resulted in differentiated management. Overall 97% and 52% of patients contributed to the per-protocol population in the control and interventions phases, respectively. The authors acknowledge this comparison may be problematic due to selection bias.

In our trial, despite non-adherence being higher in the control group, there was no apparent negative consequences on treatment outcomes or recurrence, which seems at odds to other studies. An analysis of the fluroquinolone treatment trials, albeit a non-randomised comparison, demonstrated a strong relationship between lower adherence and increased risk of recurrence(3, 13). This might mean that in the control group levels of treatment adherence were still adequate to result in the majority of patients being effectively treated and with no increased risk of recurrence(14). With the granular adherence data generated by the trial it will be important to identify whether certain patterns of non-adherence are associated with increased risk of poor end of treatment outcomes or treatment recurrence.

Alternatively, we may have underestimated recurrence, though unlikely to be differential by study group. We followed up patients for 12 months after the end of treatment, which would likely capture the vast majority of relapses(15). Recurrence in this trial, however, was very low (1·9% over 12 months), in particular compared with the fluroquinoline trials where 12-month recurrence was 2-3-fold higher. We used solid culture in laboratories that had quality control assessed prior to, though not during, the study. Sputum specimens were only collected at two time points post-end of treatment, limiting the measurement of recurrence, as well as specimens may be of lower quality compared with those collected as part of a treatment trial. Our approach for documenting recurrence, therefore, may not have been sufficiently sensitive, though we did supplement these specimens with chest radiograph.

Our trial did demonstrate a reduction in non-adherence in the intervention versus control, similar to our previous study(6), indicating improved quality of treatment with the intervention. Based on these two pragmatic trials and costing data, which suggests similar costs to routine care, the China National Tuberculosis Program has planned to expand the utilization of the monitors with real-time functions, nationwide, in their 14th five-year plan (2021-2025).

Our trial has many strengths including a large sample size, the intervention implemented by National Tuberculosis Program rather than a research team in parallel, conducted across varied settings and included follow-up of patients 12-months post end of treatment. The study does, however, have several limitations: more intensive management activities such as home visits to patients identified as having adherence problems appeared not to happen as planned; post-randomisation, two intervention clusters were combined though had minimal impact on power; use of solid rather than the more sensitive liquid culture to measure recurrence; and adherence outcomes were defined using a box-opening as synonymous with dose-taken.

Monthly review of adherence data was not adequate to influence poor treatment outcomes, in particular losses-to-follow-up. More frequent adherence data review coupled with a streamlined approach for identifying patients with adherence issues and escalating supportive management of these patients, may be successful for improving outcomes, though not supported by this study. In the real-world treatment adherence may be substandard and clinicians lack accurate methods to measure dose-taking. Digital technologies hold promise to overcome these barriers to care. The WHO uses treatment success as an indicator for performance of TB programs, though it is poor indicator of care; a patient taking less than 80% of doses may still have their outcome reported as treatment completion. Pragmatic trials of digital adherence technologies, rather than relying solely on end of treatment outcomes, could be evaluated using a combined endpoint of adherence and treatment outcome, where adherence is measured in all study groups. Recently published trials have demonstrated that digital adherence technologies can reduce lost-to-follow-up though it would be important to understand the cost of delivering such an intervention and also how these are implemented in routine practice. It is important that future trials do measure end of treatment outcomes incorporating quality treatment completion and possibly recurrence, if measured robustly, to generate strong evidence to influence policy.

## Supporting information

supplemental results

## Data Availability

All data in the present study will be available online at DataCompass (https://datacompass.lshtm.ac.uk/) on publication of the article in a peer-reviewed journal.

## Contributors

The study was conceptualized and designed by XL, BT, SH, SJ, AV, KF and YZ. XL, HD, XL, YY, XW, WH, CX, DH, HZ, SJ, YZ were responsible for data collection. JT, HD, KF were responsible for the analysis. All authors contributed to interpretation, writing of the manuscript and have approved the final submitted version of the manuscript. The corresponding author attests that all listed authors meet authorship criteria and that no others meeting the criteria have been omitted.

## Data sharing

Individual de-identified participant data that underlie results reported in this article, data dictionary, study protocol, statistical analysis plan and Stata code will be made available on LSHTM Data Compass, without restriction, immediately following publication.

## Acknowledgments

The trial was funded by a grant from the Bill & Melinda Gates Foundation (OPP1137180). China National Health Commission -Gates Foundation TB Project National Program Management Office, Beijing China - Yongxin Gao, Lili Cheng, Fanfan Zheng, Fei Huang

James J Lewis (London School of Hygiene & Tropical Medicine, London, UK)

Trial Steering Committee – Daniel Chin (Gates Foundation), Lixia Wang (Chinese Center for Disease Control and Prevention, Beijing, China), Xiexiu Wang (Chinese Anti-tuberculosis Association, Beijing China)

Endpoint Review Committee - Mengqiu Gao, Liqun Zhang, Liping Ma (Beijing Tuberculosis and Thoracic Tumor Research Institute Beijing China)

