## supplemental methods for "Digital adherence technologies to improve tuberculosis treatment outcomes: a cluster-randomised superiority trial"

### Effectiveness methods

#### 1.1. Randomisation

The restriction criteria were defined as follows:

1. There are four prefectures (six in Hangzhou, seven in Weizhou, four in Jilin and seven in Ganzhou) in three provinces (13 in Zhejiang, 4 in Jilin and 7 in Jiangxi). The difference in the number of intervention and control group clusters should be at most one in each prefecture (or province).

2. There are 14 clusters with a designated hospital and 10 with a TB dispensary. There should be equal numbers of clusters with a designated hospital in each study group (and therefore equal numbers with a TB dispensary).

3. There are 7 urban clusters and 17 rural clusters. The difference in the number of intervention and control group clusters should be at most one in each of the urban and rural clusters.

4. The difference in the average number of smear-positive TB cases notified by the TB clinic in 2015 in each cluster between intervention and control group clusters is at most 10 cases.

The process for restricted randomization followed three steps:

1. Generate 10,000 random allocations and determine the “restriction factor” for each of the four criteria above. So long as at least 1% of allocations are eligible (giving at least 27,042 eligible allocations) then proceed to step 2. 1.24% of allocations were eligible.

2. If no restrictions or stratifications are applied, the probability that any two clusters are in the same group should be 11/23 = 0.478. To ensure that valid statistical inferences can be drawn, we want to ensure that the probability that any two clusters are in the same group is not too far from 0. 478. For example, if the probability that two clusters are in the same study group is close to 1, then these two clusters may need to be treated as one cluster. Hence, the next step is to check these probabilities for all pairwise combinations (23*22/2 = 253 possibilities). This was done by simulating 5,000 acceptable allocations and then estimating the pairwise probabilities from these 5,000 allocations. The proportion where a pair of clusters were in the same study group varied between 0.288 and 0.745, which suggests that the most restrictive criteria still allows for appropriate statistical inference. Hence, these criteria were finalised for the randomisation.

3. One of the 5,000 can be chosen as the final allocation of the 24 clusters to the two groups. This was chosen using a random number generated by Stata. The random number seed to determine this was selected before any of the above steps were initiated.

All clusters were enrolled before this process began. Randomisation was conducted by James Lewis, the trial statistician at the time.

#### 1.2. Statistical methods

All analyses used a cluster-level analysis containing all clusters. For this, the outcomes were summarised to a mean, proportion, or rate for each cluster. Our primary comparison for binary and rate outcomes were ratios. To compare the study groups, we look logarithms of the cluster summaries and compared the study groups using a t-test. The exponential of the mean difference in log-summaries gave a risk or rate ratio. For the absolute difference between the study groups, the clusters summaries were compared without taking logarithms. Adjustment was done using a two stage approach. First, a model was fit with the outcome ad the dependent variable and adjustment covariates as the independent variables ignoring the intervention and clustering. This provided estimated residuals, which were summarised by cluster and analysed as described above.

Multiple imputation

For the primary composite outcome participants were classified as having a favourable outcome if: they had a treatment outcome at the end of treatment of cured or completed treatment and no recurrence defined by either a negative culture at 18 months or at 18 months, culture is missing, or the patient was unable to produce sputum and they have no signs of new active TB meaning (1) no self-reported restart of treatment or (2) no sign of new active TB on x-ray. The composite outcome was missing if a participant meets neither definition of a favourable or unfavourable outcome, particularly an issue if a patient has become lost to follow up after treatment completion, with no sign of a recurrence at their last follow-up visit.

For the primary outcome, multiple imputation was used to prevent bias arising from lost to follow up after completion of treatment. Chained equations were used to imputed poor treatment outcome, recurrence up to 12 months, and recurrence up to 18 months. Recurrence was defined by a positive culture at or before the designated time point, an x-ray indicative of TB, or a self-report of restarting treatment. Non-recurrence required a negative culture at the designated time. Otherwise, recurrence was missing and imputed. Components were simplified to good or poor treatment outcome, recurrence up to 12 months, or recurrence between 12 and 18 months. Imputations were adjusted for cluster and each of the other components with data augmentation to overcome perfect prediction. Imputation models included all adjustment covariates, cluster, and the two other components of the model. It was pre-specified that problematic covariates would be removed from the imputation. This resulted in a final set of imputation models that contained only cluster, and the other two component of a poor outcome.

Complete case analysis

For the primary outcome, as a sensitivity analysis, a complete case analysis was also conducted. For all other outcomes, complete case analysis was the primary analysis. These analyses only included patients with a definable outcome.

For the primary composite outcome, classification of no TB recurrence was loosened so that a self-report of no restart of treatment, or a chest x-ray showing no signs of active TB at 18 months were sufficient for a patient to be classified as having a good composite outcome.

Subgroup analyses

For individual level characteristics, first calculate the risk of a poor outcome in each subgroup and cluster. Calculate the mean of these risks within subgroup to estimate the risk of a poor outcome in control and intervention groups. To test for a difference, calculate the difference between the two subgroups within each cluster. A t-test is performed on these differences to test for effect modification.

For cluster level characteristics, a linear regression will be used with study group, subgroup, and an interaction between the two as independent variables. A likelihood ratio test will be used to estimate a p-value for the interaction. The risk of a poor outcome and its confidence interval in each group will be calculated from the relevant coefficients from the regression.
