## supplemental results for "Digital adherence technologies to improve tuberculosis treatment outcomes: a cluster-randomised superiority trial"

### Effectiveness Results

#### 1.1. Characteristics of the trial sites and participants

Early in the trial, 19 patients were incorrectly counted as post-enrolment exclusions after enrolment due to misunderstandings of the trial protocols. Additional training was provided to trial sites, and these have been included in the intention to treat population despite missing values for most outcomes.

#### 1.2. Per-protocol population

94.7% (1231/1300) and 93.4% (1156/1238) contributed to the per-protocol complete case analysis for the primary outcome, in the control (routine care) and intervention groups, respectively.

#### 1.3. Process measures

The rate of medication event reminder monitor (MERM) malfunctions or errors was similar between the groups (0.2 and 0.4 days of treatment affected by errors per person-month in control and intervention groups respectively; adjusted rate ratio=1.75[0.91,3.36]). Patients in the intervention group were more likely to report non-use of the MERM due to travel (intervention arm 1.8% of days not using the intervention vs control arm 0.9% of days not using the MERM). Intervention group patients were 1.30(1.06, 1.60) time more likely than control patients to open the box for a short time that would not be long enough for this to signify taking treatment (<2 seconds). Length of treatment was similar between the groups (mean 6 months control vs 6.1 months intervention, adjusted mean difference=+0.1[-0.3,+0.3]).

There was no difference between the groups in withdrawal from use of the MERM (control 105[GM 6%] vs intervention 106[GM 8%]); most of these were due to inability to use fixed dose combination therapy, which was also similar between the groups (control 79[GM 5%] vs intervention 46[GM 4%]).
