## supplemental figure 1 for "Digital adherence technologies to improve tuberculosis treatment outcomes: a cluster-randomised superiority trial"

Figure S1: Adherence to treatment (measured by MERM) by month of treatment and study group

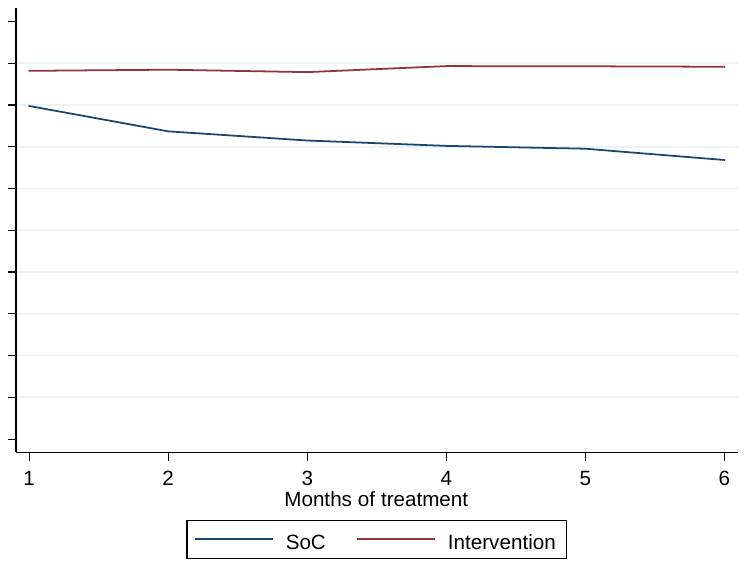

SoC standard of care (control)
