## Supplementary figures and images for "Digital adherence technologies to improve tuberculosis treatment outcomes: a cluster-randomised superiority trial"

### supplemental figure 2

Figure S2: Self-reported supervisor of medication, control arm

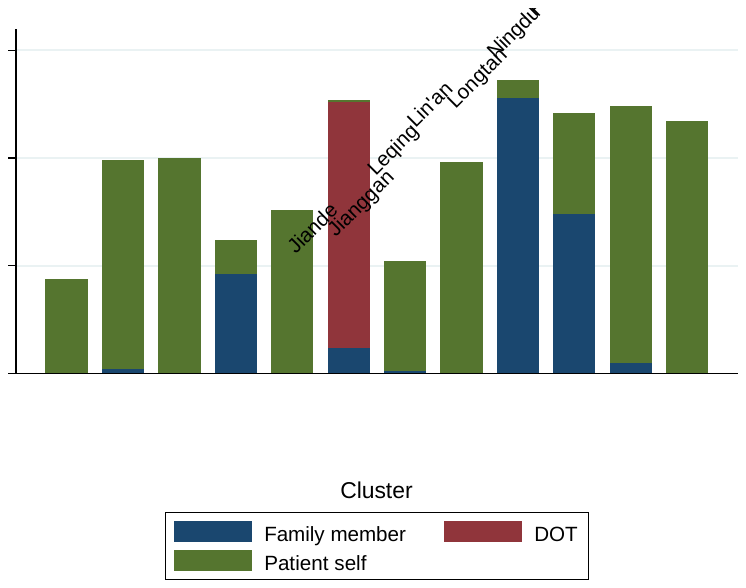

DOT directly observed therapy
