## supplemental table 1 for "Digital adherence technologies to improve tuberculosis treatment outcomes: a cluster-randomised superiority trial"

Table S1: Additional baseline characteristics of clusters

|  | Control | Intervention |
| --- | --- | --- |
| Cluster level covariate | N=12 | N=11 |
| Number of townships, median(IQR) | 17.5 (13.5-24.5) | 21 (14-25) |
| Number of villages, median(IQR) | 289.5(190-451.5) | 257(136-520) |
| Number of doctors seeing TB patients, median (IQR) | 2.5 (1.5-4) | 2(1-3) |
| Per capita net income (CYN), median(IQR) | 27877.5  (13653-38439) | 34087  (17232.5-39238) |
| Population size per 1000, median(IQR) | 603.5 (497.0-859.4) | 760.0 (439.2-1084.8) |
| Laboratory onsite, n(%) | 12 (100%) | 11 (100%) |
| X-ray onsite, n(%) | 12 (100%) | 11 (100%) |
| 1 month of TB medication dispensed during intensive phase, n(%) | 12 (100%) | 11 (100%) |
| 1 month of TB medication dispensed during continuation phase, n(%) | 12 (100%) | 11 (100%) |
| Are incentives offered, n(%) | 2 (17%) | 1 (9%) |
| Individual level covariate | **N = 1388** | **N=1298** |
| Number of household members, median (IQR) | 4 (3 – 5) | 4 (3 - 5) |
| Have sufficient money to cover costs, n(%) | 1099 (79%) | 1147 (88%) |
| Distance to TB clinic, median (IQR) | 15 (5 – 30) | 15 (5 – 28) |
| Distance to supervision facility, median (IQR) | 2 (1 – 3) | 2 (1 – 2.5) |
| IQR interquartile range; CNY Chinese Yuan; X-ray chest radiograph | | |
