## supplemental table 2 for "Digital adherence technologies to improve tuberculosis treatment outcomes: a cluster-randomised superiority trial"

Table S2: Summary of the primary composite outcome and components of the unfavourable outcome, by group

|  | Control |  | Intervention |  |
| --- | --- | --- | --- | --- |
|  | **n** | **%** | **n (%)** |  |
| Total number of participants | **1388** |  | **1298** |  |
| Composite outcome missing (undefined) | 88 | 6.3% | 60 | 4.6% |
| Composite outcome defined: | **1300** |  | **1238** |  |
| Composite favourable outcome^a^ | **1083** | **83.3%** | **1022** | **82.6%** |
| Composite unfavourable outcome^a^ | **217** | **16.7%** | **216** | **17.4%** |
| *Poor end of treatment outcome^b^:* | *203* | *93.5%* | *188* | *87.0%* |
| Lost to follow up during treatment | 156 |  | 133 |  |
| Treatment failure | 33 |  | 45 |  |
| Death (TB or non-TB) | 12 |  | 10 |  |
| Switch to MDR | 2 |  | 0 |  |
| *Recurrence in those with a good treatment outcome:* | *14* | *6.5%* | *28* | *13.0%* |
| Positive culture | 9 |  | 22 |  |
| X-ray indicative of active TB only | 1 |  | 2 |  |
| TB retreatment only | 2 |  | 3 |  |
| X-ray indicative of active TB and TB retreatment | 2 |  | 1 |  |

^a^ percentage of those with outcome defined; ^b^ percentage of those with a composite unfavourable outcome; X-ray chest radiograph
