## supplemental table 3 for "Digital adherence technologies to improve tuberculosis treatment outcomes: a cluster-randomised superiority trial"

Table S3: Cluster summaries of primary unfavourable outcome (poor end of treatment outcome or recurrence)

| Control |  | Intervention |  |
| --- | --- | --- | --- |
| Mean of imputations (%) | Complete case n/N(%) | Mean of imputations (%) | Complete case n/N(%) |
| 8.2 | 6/93 (6.4) | 8.3 | 11/138 (8.0) |
| 9.5 | 8/82 (9.8) | 9.1 | 7/80 (8.8) |
| 11.9 | 12/106 (11.3) | 10.2 | 8/79 (10.1) |
| 13.0 | 17/134 (12.7) | 11.7 | 14/114 (12.3) |
| 15.4 | 22/137 (16.1) | 15.8 | 23/144 (16.0) |
| 17.0 | 11/64 (17.2) | 17.4 | 25/142 (17.6) |
| 18.5 | 26/147 (17.7) | 19.5 | 24/120 (20.0) |
| 19.0 | 29/153 (19.0) | 19.9 | 30/150 (20.0) |
| 22.4 | 33/151 (21.8) | 22.4 | 16/72 (22.2) |
| 22.9 | 10/53 (18.9) | 28.6 | 44/155 (28.4) |
| 23.4 | 24/115 (20.9) | 31.5 | 14/44 (31.8) |
| 29.1 | 19/65 (29.2) |  |  |
