## supplemental table 4 for "Digital adherence technologies to improve tuberculosis treatment outcomes: a cluster-randomised superiority trial"

Table S4: pre-specified and post-hoc sensitivity analyses for the primary unfavourable outcome

|  | Control | Intervention | Unadjusted arm comparison (95% CI) | Adjusted arm comparison (95% CI) |
| --- | --- | --- | --- | --- |
| Risk difference ^a^ |  |  |  |  |
| Multiple imputation | 239/1388 (18%) | 224/1298 (18%) | 0.2% (-6.4%, 6.7%) | 0.7% (-4.5%, 5.9%) |
| Complete case unfavourable outcome | 217/1300  (17%) | 216/1238 (18%) | 1.0% (-5.1%, 7.0%) | 1.6% (-3.1%, 6.2%) |
| Risk ratio (complete case, pre-specified, sensitivity analyses) |  |  |  |  |
| Intention to treat | 217/1300 (16%) | 216/1238 (16%) | 1.03 (0.71, 1.51) | 1.05 (0.78, 1.41) |
| Per-protocol | 205/1231 (15%) | 195/1156 (16%) | 1.04 (0.69, 1.57) | 1.06 (0.78, 1.45) |
| Risk ratio (Post-hoc sensitivity analyses) |  |  |  |  |
| Excluding NTM | 217/1300 (16%) | 215/1237 (16%) | 1.03 (0.71, 1.51) | 1.05 (0.78, 1.40) |
| Setting treatment 9 months or long as poor outcome ^b^ | 255/1304 (19%) | 246/1242 (19%) | 1.01 (0.74, 1.38) | 1.02 (0.76, 1.36) |
| Setting unable to produce sputum as negative culture in multiple imputation ^b^ | 239/1388 (16%) | 224/1298 (16%) | 0.99 (0.66, 1.48) | 1.01 (0.73, 1.40) |
| All sensitivity analysis used complete cases and unless specified used intention to treat populations and were pre-specified in the analysis plan  ^a^ Percentages shown are arithmetic means of the cluster level risks  ^b^ Post-hoc: Not pre-specified in analysis plan  NTM Non‐tuberculous mycobacteria; CI confidence interval. | | | | |
