## supplemental table 5 for "Digital adherence technologies to improve tuberculosis treatment outcomes: a cluster-randomised superiority trial"

Table S5: Sensitivity analyses for secondary treatment adherence outcomes

|  | Control | Intervention | Unadjusted risk ratio (95% CI) | Adjusted risk ratio (95% CI) |
| --- | --- | --- | --- | --- |
| Months in which patients missed >20% of doses including days patients reported not using MERM / Months of treatment per person (%) | 2.7/6.0 (47%) | 1.1/6.0 (19%) | 0.42 (0.31, 0.57) | 0.44 (0.34, 0.56) |
| Doses missed including days patients reported not using MERM mean /doses expected per person (%) | 45/163 (28%) | 20/165 (13%) | 0.46 (0.36, 0.58) | 0.48 (0.40, 0.59) |
| Post-hoc |  |  |  |  |
| Months in which patients missed >20% of doses including LFTU as non-adherence / Months of treatment per person(%) | 3.0 / 6.2 (48%) | 1.2/6.2 (20%) | 0.41 (0.30, 0.57) | 0.43 (0.33, 0.56) |
| Doses missed including LTFU as non-adherence / Doses expected per person(%) | 48/164 (30%) | 23/165(15%) | 0.50 (0.39, 0.64) | 0.52 (0.42, 0.65) |

MERM medication event reminder monitor; LTFU lost to follow-up

Adherence outcomes summarised by patient, taking the arithmetic mean within cluster, then the geometric mean between clusters
