## supplemental table 6 for "Digital adherence technologies to improve tuberculosis treatment outcomes: a cluster-randomised superiority trial"

Table S6: Post-hoc adherence measures

|  | Control | Intervention |
| --- | --- | --- |
| Patients that missed at least one dose n/N (%) | 1300/1306 (>99%) | 1244/1261 (99%) |
| Number of times doses missed per patient mean (sd) | 44.9 (39.3) | 17.4 (16.1) |
| Days to first missed dose mean (sd) | 87.6 (55.8) | 87.1 (56.5) |
| Patients that ever missed 2 or more consecutive doses n/N (%) | 1149/1306 (88%) | 870/1261 (69%) |
| Number of times patients missed 2 or more consecutive doses mean(sd) | 7.3 (7.1) | 2.5 (3.4) |
| Patients that ever missed 3 or more consecutive doses n/N (%) | 987/1306 (76%) | 471/1261 (37%) |
| Number of times patients missed 3 or more consecutive doses mean(sd) | 4.1 (4.9) | 0.8 (1.7) |
| Patients that ever missed 4 or more consecutive doses n/N (%) | 828/1306 (63%) | 296/1261 (23%) |
| Number of times patients missed 4 or more consecutive doses mean(sd) | 2.5 (3.4) | 0.4 (1.0) |

Analysis ignores missing data in counting consecutive doses. Similar result were found if missing data prevented doses being considered consecutive. sd standard deviation
