## supplemental table 7 for "Digital adherence technologies to improve tuberculosis treatment outcomes: a cluster-randomised superiority trial"

Table S7: Additional process measures

|  | Control | Intervention |
| --- | --- | --- |
| N | 1351 | 1283 |
| Months of treatment, mean (sd) ^a^ | 6.1 (0.4) | 6.1 (0.3) |
| Patient ever receive > 1 months of drugs with agreement of doctor n/ total n (%^b^) | 265/1387 (6%) | 89/1296 (4%) |
| [Intervention only] |  |  |
| Times alarm sounded before opening, Mean (sd), N |  | 1.4 (0.1), 1261 |
| Required intensive management n/total n(%^b^) |  | 196/1261 (8%) |
| Received intensive management |  | 156/190 (74%) |
| Visits from township/village doctor per month after change in management mean (sd) |  | 1.8 (1.0) |
| Required DOT n/total n(%^b^) |  | 100/1261 (4%) |
| Received DOT n/total n(%^b^) |  | 53/99 **^c^** (51%) |
| ^a^ Unadjusted mean difference: 0.0 (-0.3, +0.4) p= 0.87; Adjusted mean difference: 0.0 (-0.3, 0.3) p=0.92.  sd standard deviation; DOT directly observed treatment.  ^b^ Geometric mean; ^c^ missing for one patient | | |
